# Impact of COVID-19 on Primary Care Mental Health Services: A Descriptive, Cross-Sectional Timeseries of Electronic Healthcare Records

**DOI:** 10.1101/2020.08.15.20175562

**Authors:** Clarissa Bauer-Staeb, Alice Davis, Theresa Smith, Wendy Wilsher, David Betts, Chris Eldridge, Emma Griffith, Julian Faraway, Katherine Button

## Abstract

**Introduction:** There are growing concerns about the impact of the COVID-19 pandemic on mental health. With government-imposed restrictions as well as a general burden on healthcare systems, the pandemic has the potential to disrupt the access to, and delivery of, mental healthcare. Ultimately, this could potentially lead to unmet needs of individuals requiring mental health support.

**Methods:** Electronic healthcare records from primary care psychological therapy services (Improving Access to Psychological Therapy) in England were used to examine changes in access to mental health services and service delivery during early stages of the COVID-19 pandemic. A cross-sectional, descriptive timeseries was conducted using data from 1^st^ January 2019 to 24^th^ May 2020 across five NHS trusts to examine patterns in referrals to services (n = 171,823) and appointments taking place (n = 865,902).

**Results:** The number of patients accessing mental health services dropped by an average of 55% in the 9 weeks after lockdown was announced, reaching a maximum reduction of 74% in the initial 3 weeks after lockdown in the UK. As referrals began to increase again, there was a relatively faster increase in referrals from Black, Asian, and ethnic minority groups as well an increase in referrals from more densely populated areas. Despite a reduction in access, service providers adapted to infection control guidance by rapidly shifting to remote delivery of care.

**Interpretation:** Services were able to rapidly adapt to provide continuity of care in mental healthcare. However, patients accessing services reduced dramatically, potentially placing a future burden on service providers to treat a likely backlog of patients in addition to a possible excess of patients as the long-term consequences of the pandemic become more apparent. Despite the observational nature of the data, which should be noted, the present study can inform the planning of service provision and policy.

## BACKGROUND

Primary care services are often the first port of call for patients with common mental health problems, with patients showing a preference for psychological therapy compared to medication (McHugh, Whitton, Peckham, Welge, & Otto, 2013). Psychological therapy for depression and anxiety in primary care settings is delivered by Improving Access to Psychological Therapy (IAPT) services in England, with approximately 1.69 million referrals to IAPT in 2019-20 (Clark, Canvin, Green, Layard, Pilling & Janecka, 2018; NHS Digital, 2020).

There is growing concern about the profound and long-lasting impacts of COVID-19 on mental health (Holmes et al., 2020). National surveys during the pandemic in the UK, examining the psychosocial impact of COVID-19, showed that 24.4% and 31.4% of people have moderate to severe symptoms of anxiety and depression, respectively – with little change over time (Fancourt, Steptoe & Bu, 2020). People who have previously or are currently suffering from mental health conditions, as well as those who become mentally unwell during the pandemic, may potentially be vulnerable groups (Holmes et al., 2020). The pandemic may also disproportionately affect the mental health of other groups, including those with physical health conditions, individuals facing financial instability, ethnic minority groups, as well as young and older adults (Bu, Steptoe, & Fancourt, 2020; Frank, Iob, Steptoe & Fancourt, 2020; Holmes et al., 2020; Moorthy & Sankar, 2020). The provision of adequate mental health support to address the psychological impact of the pandemic and meet mental health needs is critical.

Despite the growing concerns about COVID-19 on mental health, less focus has been placed on how individuals with mental health problems are supported (Johnson et al., 2020). Concerns have been raised about adequate service provision during the pandemic (Holmes et al., 2020). However, early research in secondary care suggests a relative stability in mental healthcare (Stewart, Martin, & Broadbent, 2020). Nonetheless, staff shortages and service reconfigurations as well as the pressures of implementing infection control measures pose challenges to mental health staff (Johnson et al., 2020). To date, very little is known about whether individuals – particularly vulnerable groups – have adequate access to mental healthcare and how the care of current patients has been impacted in primary care settings.

The use of electronic healthcare records provides a first avenue to examine the impact of COVID-19 on primary care mental health services at scale (Holmes et al., 2020). Using electronic healthcare records from IAPT services, we aim to understand service use, as well as examine trends in who is accessing services and how they are being accessed. Furthermore, we aim to understand the impact of COVID-19 on service delivery and how service providers adapted during the early stages of the pandemic.

## METHODS

### Settings

Improving Access to Psychological Therapies (IAPT) are primary care services in England delivering psychological interventions for depression and anxiety (Clark, et al., 2018). A minimum dataset is routinely collected for all patients, recording data relating to their demographic and clinical characteristics as well as their treatment (Clark, et al., 2018).

### Design

The present study is cross-sectional, examining all referrals and appointments in five NHS trusts across the south of England from 1^st^ January 2019 until 24^th^ May 2020. The dataset contained information for 64 weeks before the UK lockdown, implemented on 23^rd^ March 2020, and the et al., subsequent 9 weeks after lockdown was announced. All data were extracted and fully anonymised by Mayden.

### Measures

To examine the impact of COVID-19 on access to services, we examined the data of all incoming referrals to IAPT. Specifically, we consider the number of referrals, referral sources, as well as sociodemographic and clinical variables of referrals including gender, ethnicity, long-term condition status, age, previous referrals to IAPT, baseline Patient Health Questionnaire-9 (PHQ-9) and baseline Generalised Anxiety Disorder Scale-7 (GAD-7) (Kroenke, Spitzer & Williams, 2001; Spitzer, Kroenke & Williams, 2006). Additionally, the Index of Multiple Deprivation (IMD) of referrals was determined as a proxy for socioeconomic status and population density (people per square kilometre) as a proxy for urbanicity (Noble et al., 2019; Park, 2020). IMD and population density were determined at the Lower Super Output Area level via linkage to the Office of National Statistics databases. To examine the impact of COVID-19 on service provision, we examined the data for all appointments, specifically the number of appointments and the consultation medium for attended appointments. Variable definitions can be found in Supplementary Material A.

### Statistical Analysis

Descriptive characteristics are presented for the entire timeframe from 1^st^ January 2019 to 24th May 2020. To retain anonymity of service providers, data is presented in aggregate across all NHS trusts. Timeseries are presented containing the weekly total count for categorical variables and weekly averages for continuous variables. To quantify changes in access, weekly counts of incoming referrals for the 9 weeks before and after lockdown were compared to the corresponding weeks in 2019.

At the patient-level, missing data for factor variables were defined as an additional factor level. Missing data for continuous variables were excluded. The frequency of missing data for IMD and population density was < 0.5%. At the patient-level, average missing data for the baseline PHQ-9 and GAD-7 was 28%. Weekly mean missing data for baseline PHQ-9 and GAD-7 was consistently < 40% throughout the year. There may be various reasons as to why no baseline measures are taken, including patients having never attended an appointment. The last week was excluded from analyses as more than 40% of the baseline PHQ-9 and GAD-7 were missing, exceeding the annual weekly maximum. This possibly reflects that people referred close to the date of data extraction were unlikely to have had a first appointment booked within this short timeframe. As such, baseline PHQ-9 and GAD-7 scores are only presented until 17^th^ May 2020. For missing continuous data at the aggregate level, the last recorded observation was carried forward to estimate weekly averages.

All analyses were performed in the R programming language (R Core Team, 2013).

## RESULTS

### Sample Characteristics

In the timeframe of 1^st^ January 2019 to 24^th^ May 2020, 171,823 referrals came into IAPT services across five NHS trusts. The majority of referrals were self-referrals, typically female, White, with an average age of 38. Throughout the same duration, 865,902 appointments were scheduled amongst which the majority were attended and took place face-to-face.

**Table 1.**
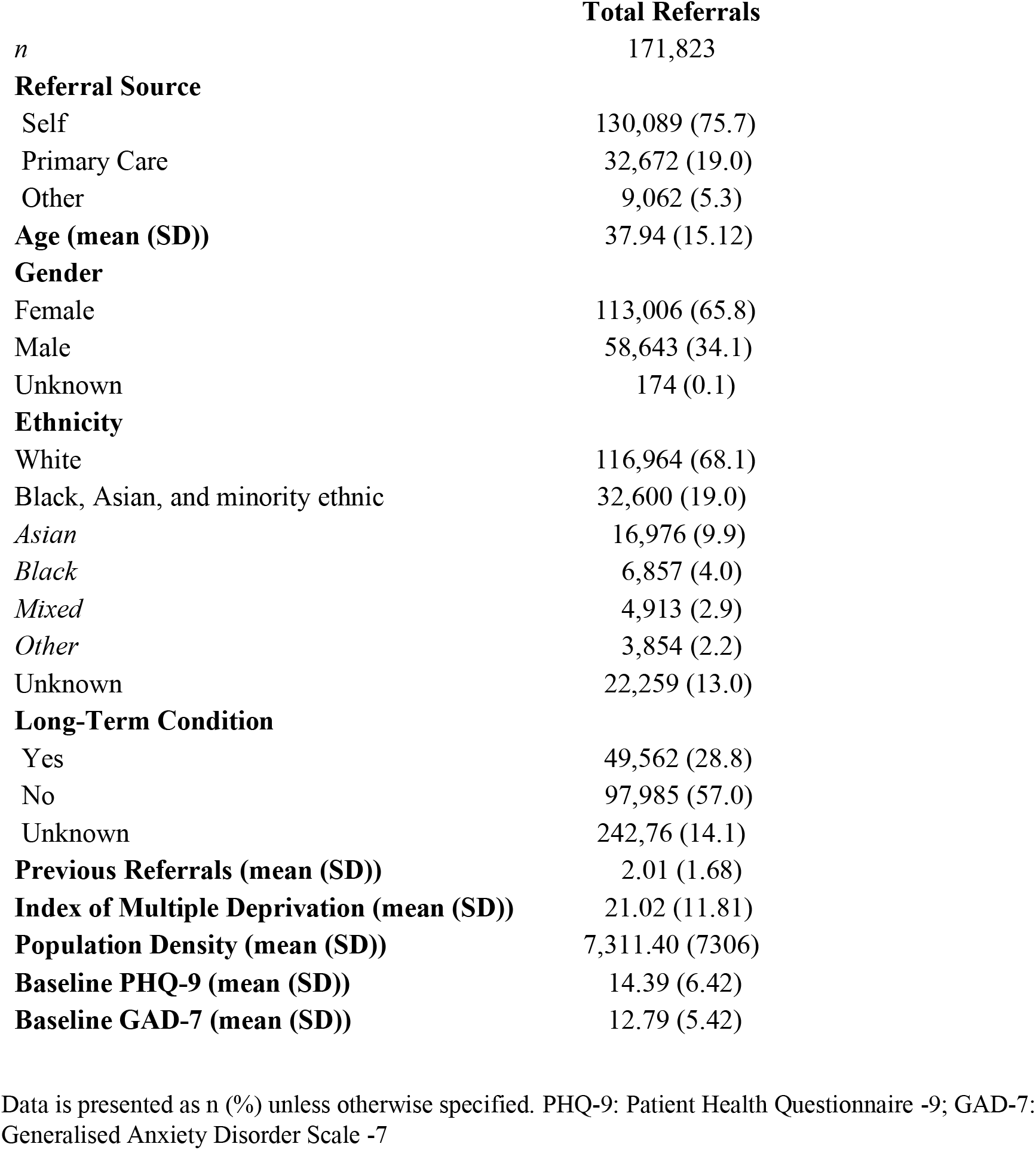
Characteristics of referrals from 1st January 2019 to 24^th^ May 2020.

**Table 2.**
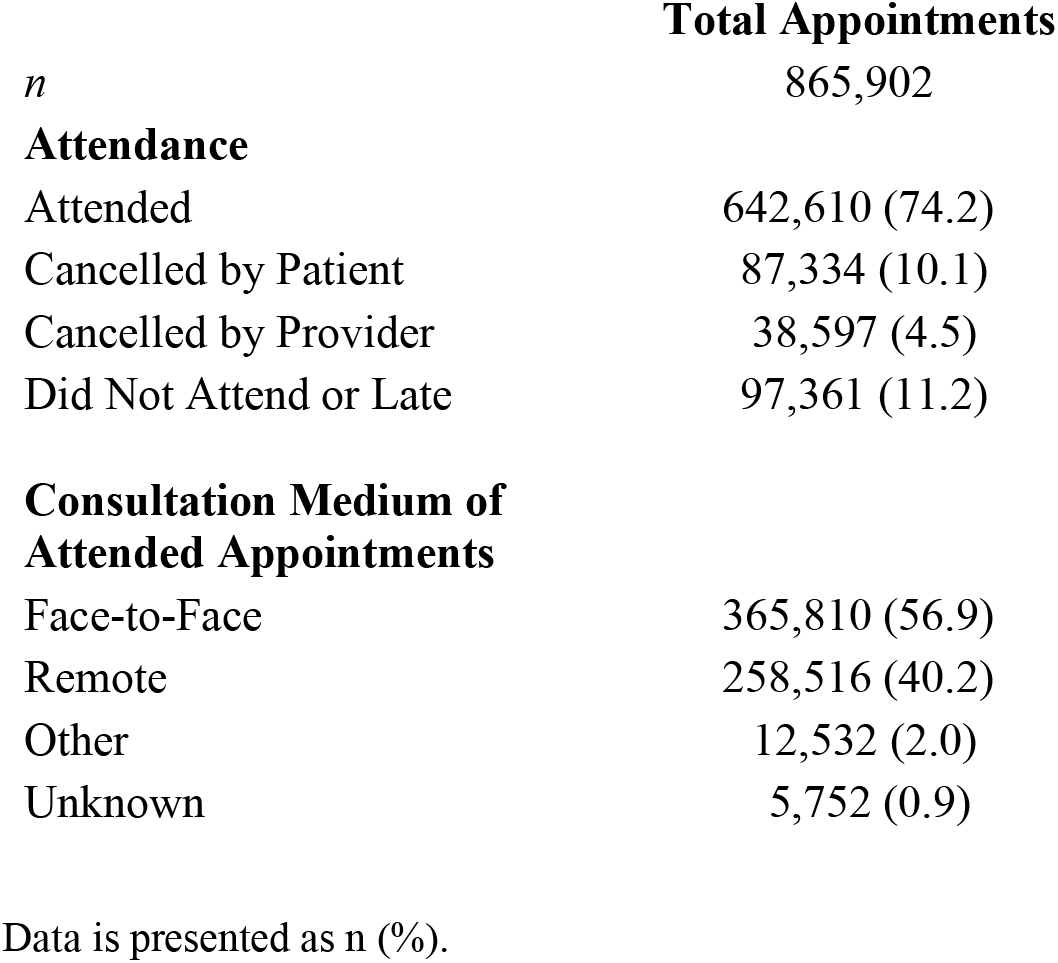
Characteristics of appointments from 1^st^ January 2019 to 24^th^ May 2020.

Figure 1 demonstrates that there was a significant decline in referrals in March 2020. The decline in referrals commenced approximately one week prior to the official government announcement of a UK-wide lockdown beginning on 23^rd^ March 2020. Table 3 shows the decline in referrals was greatest in the immediate three weeks after lockdown, reaching a maximum reduction of 74% in referrals compared to the same time in 2019. The decline in referrals is of a similar magnitude to a decline observed at the end of December during the Christmas holidays, albeit slightly larger.

**Figure 1.**
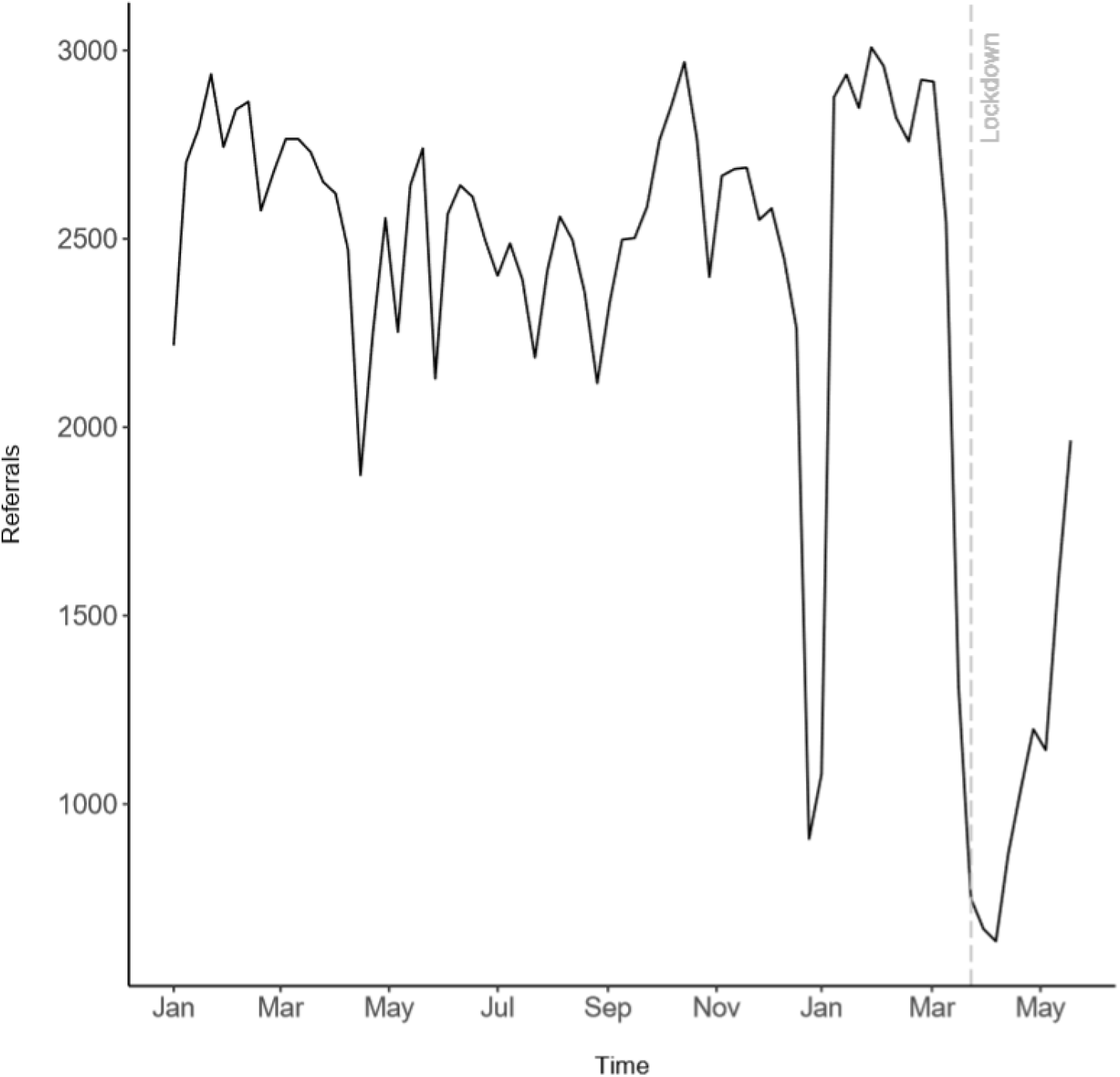
Total weekly referrals from 1st January 2019 to the 24^th^ May 2020

**Table 3.** Total weekly referrals 9 weeks before and after lockdown compared between 2019 and 2020.

|  | **Referrals 2019** | **Referrals 2020** | **Difference (n)** | **Difference (%)** |
| --- | --- | --- | --- | --- |
|  | 2,937 | 2,847 | -90 | -3 |
|  | 2,744 | 3,008 | 264 | 10 |
|  | 2,843 | 2,958 | 115 | 4 |
|  | 2,864 | 2,821 | -43 | -2 |
|  | 2,575 | 2,758 | 183 | 7 |
|  | 2,675 | 2,922 | 247 | 9 |
|  | 2,765 | 2,917 | 152 | 5 |
|  | 2,765 | 2,545 | -220 | -8 |
|  | 2,730 | 1,312 | -1,418 | -52 |
|  | 2,651 | 749 | -1,902 | -72 |
|  | 2,620 | 669 | -1,951 | -74 |
|  | 2,471 | 636 | -1,835 | -74 |
|  | 1,873 | 870 | -1,003 | -54 |
|  | 2,253 | 1,040 | -1,213 | -54 |
|  | 2,555 | 1,199 | -1,356 | -53 |
|  | 2,253 | 1,143 | -1,110 | -49 |
|  | 2,642 | 1,588 | -1,054 | -40 |
|  | 2,740 | 1,965 | -775 | -28 |
| **Total after**<br>**Lockdown** | 22,058 | 9,859 | -12,199 | -55 |

Solid line denotes lockdown

In the fourth week after the lockdown was announced, referrals started to gradually increase again over time. However, referrals had not fully recovered by the end of May. The total number of referrals in the week commencing on 18^th^ May were still 28% lower than the corresponding week in 2019.

In the 9 weeks after the UK entered lockdown, there was an average 55% reduction in referrals compared to the corresponding weeks in 2019. In the present dataset, this translated into 12,199 fewer patients accessing mental health services than might be expected for that time of year.

**Figure 2.**
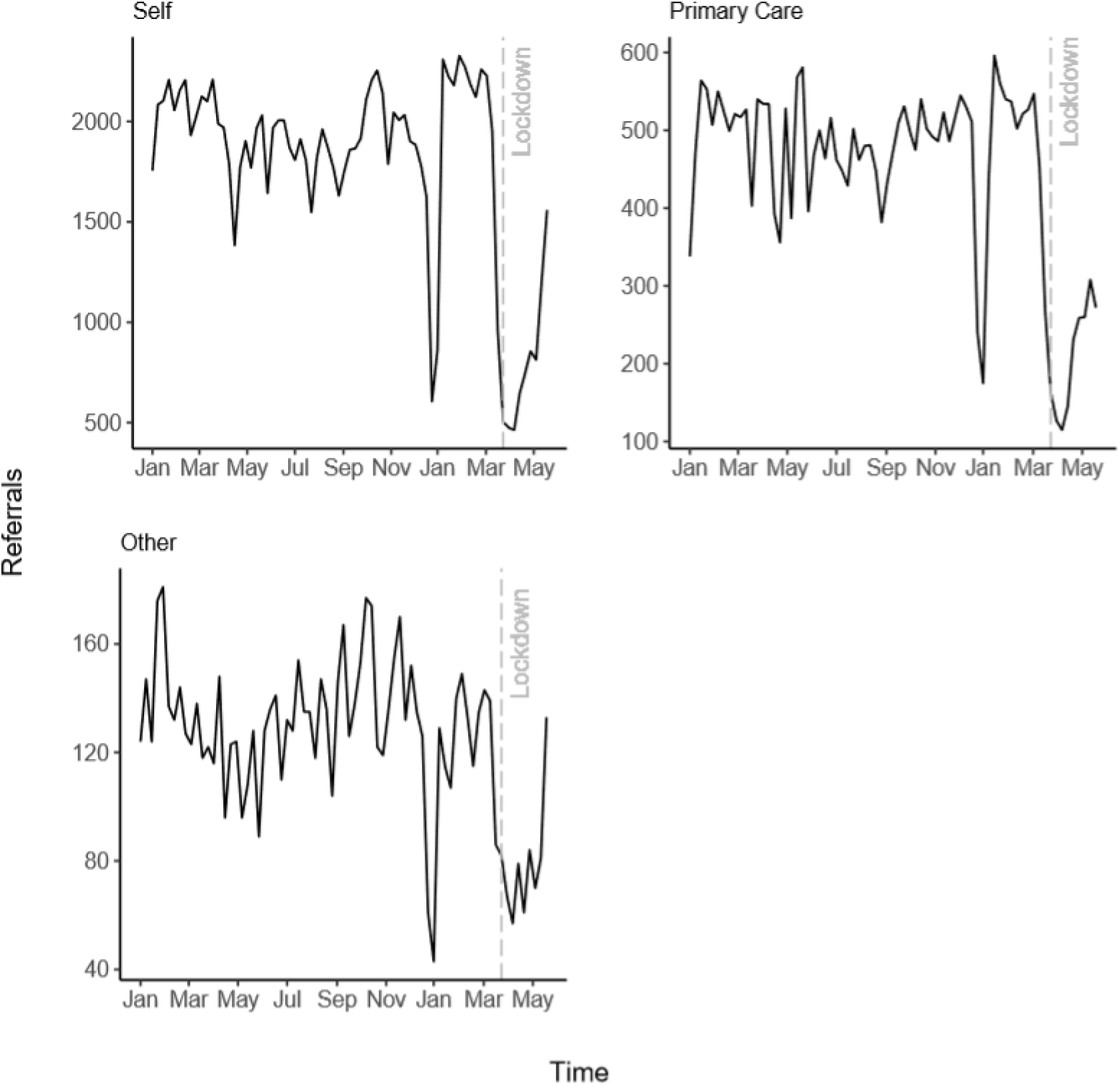
Total weekly referrals by referral source from 1st January 2019 to 24th May 2020

There was a reduction in referrals across all sources after the lockdown was imposed, with self-referrals and referrals from other sources returning to baseline fastest, while referrals from primary care were increasing at a slower rate.

### Sociodemographic characteristics of referrals

There are no clear changes in referrals by gender, age, long-term condition status or mean number of previous referrals (Supplementary Material B).

**Figure 3.**
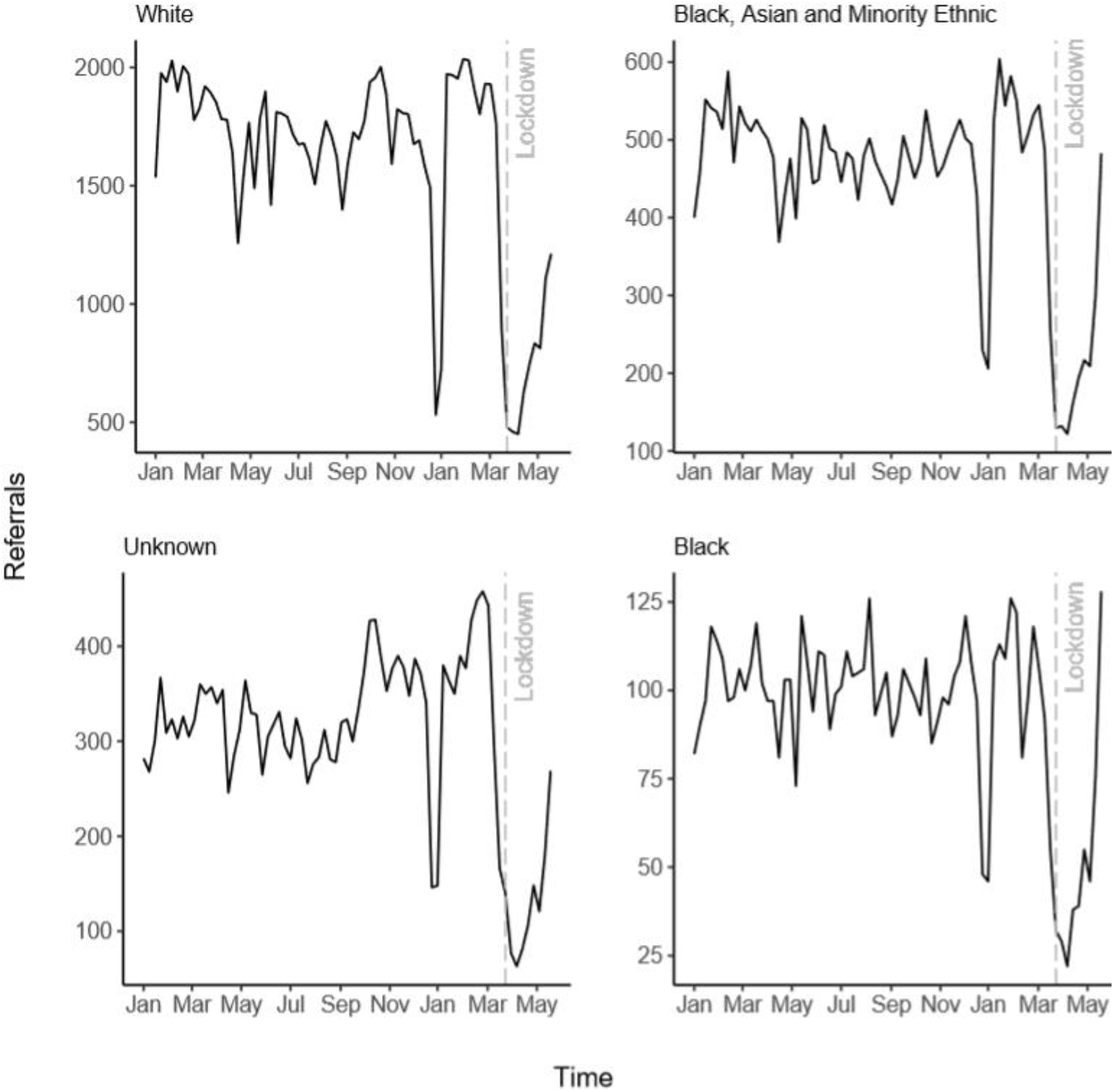
Total weekly referrals by ethnicity from 1st January 2019 to 24^th^ May 2020

Compared to referrals from a White background, Black, Asian and minority ethnic (BAME) referrals appeared to increase again at a faster rate after the initial drop observed around lockdown, being approximately equal to the corresponding timepoint in 2019. When examining BAME subgroups, there was a particular increase in referrals towards the end of May by patients with a Black ethnic background, reaching the highest number of referrals observed across the entire timespan.

**Figure 4.**
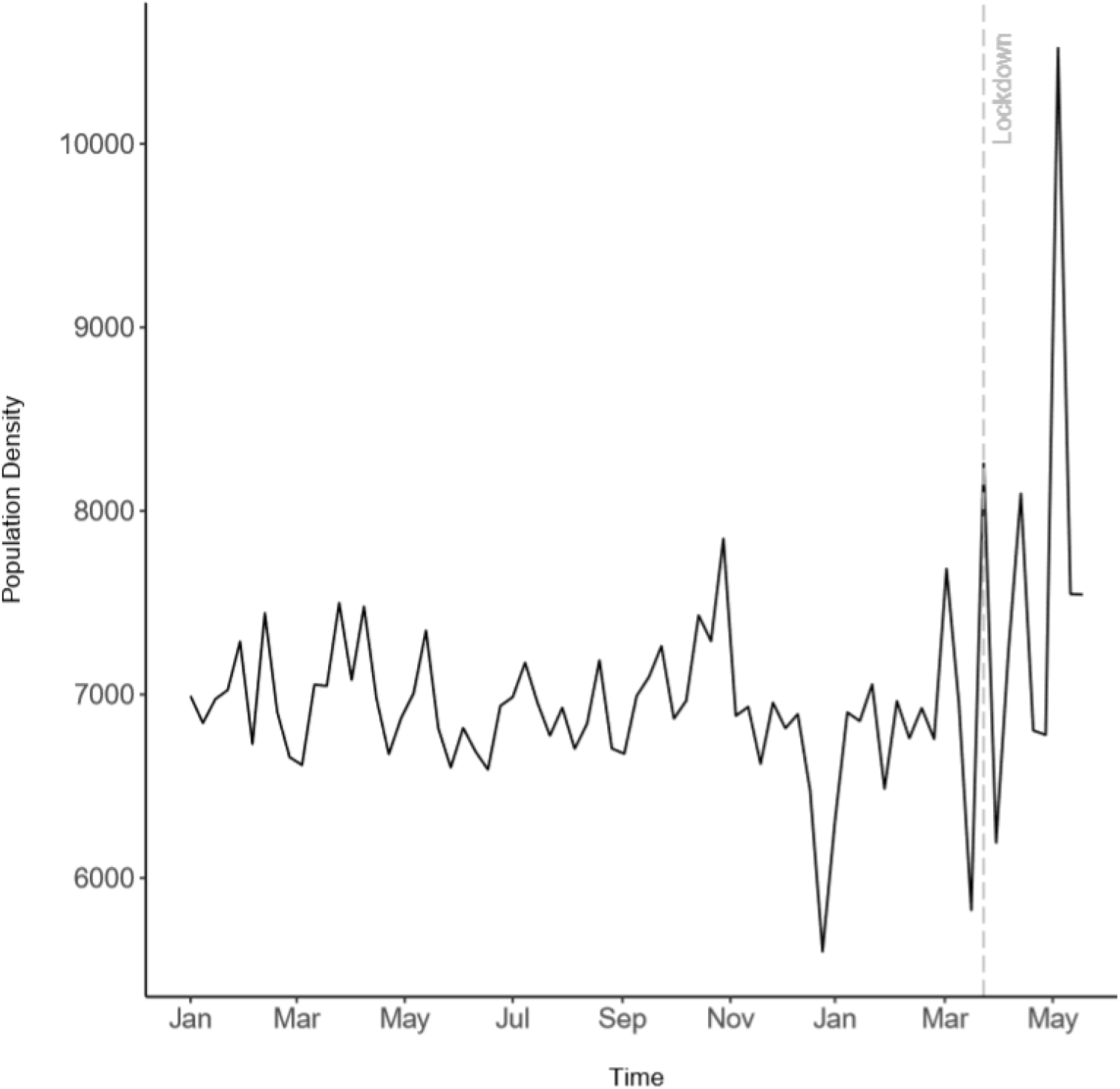
Weekly average population density of referrals from 1st January 2019 to 24th May 2020

There appears to be an increase in referrals from areas with higher population densities after lockdown was imposed.

### Baseline Depression and Anxiety of Referrals

**Figure 5.**
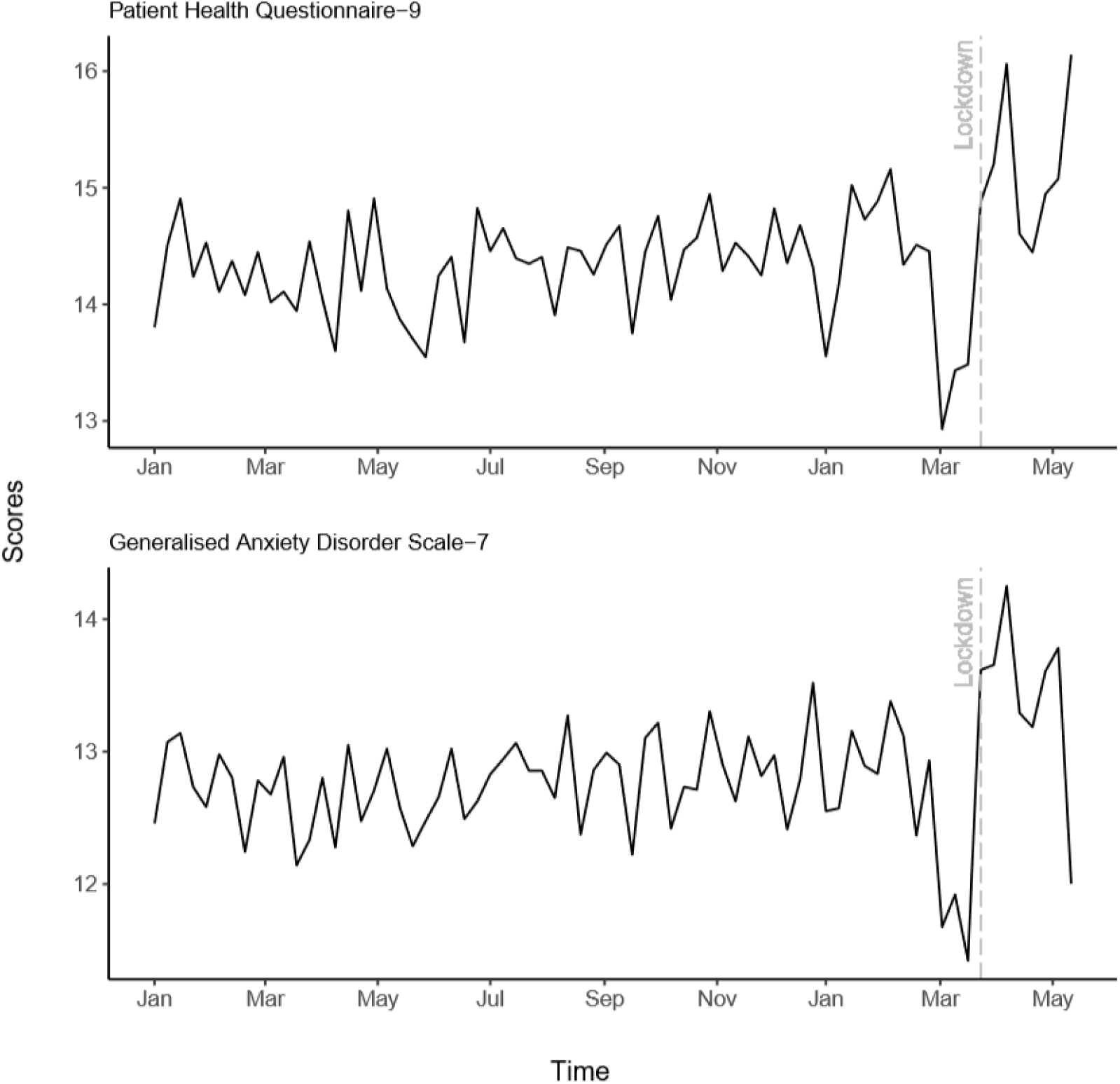
Weekly average baseline depression and anxiety scores for incoming referrals from 1st January 2019 to 17^th^ May 2020

There appears to be a reduction in average weekly first PHQ-9 and GAD-7 scores prior to lockdown, with an increase immediately after lockdown was imposed. This increase appears to be sustained throughout May for the PHQ-9, with the first GAD-7 scores returning to baseline levels.

### Appointments

**Figure 6.**
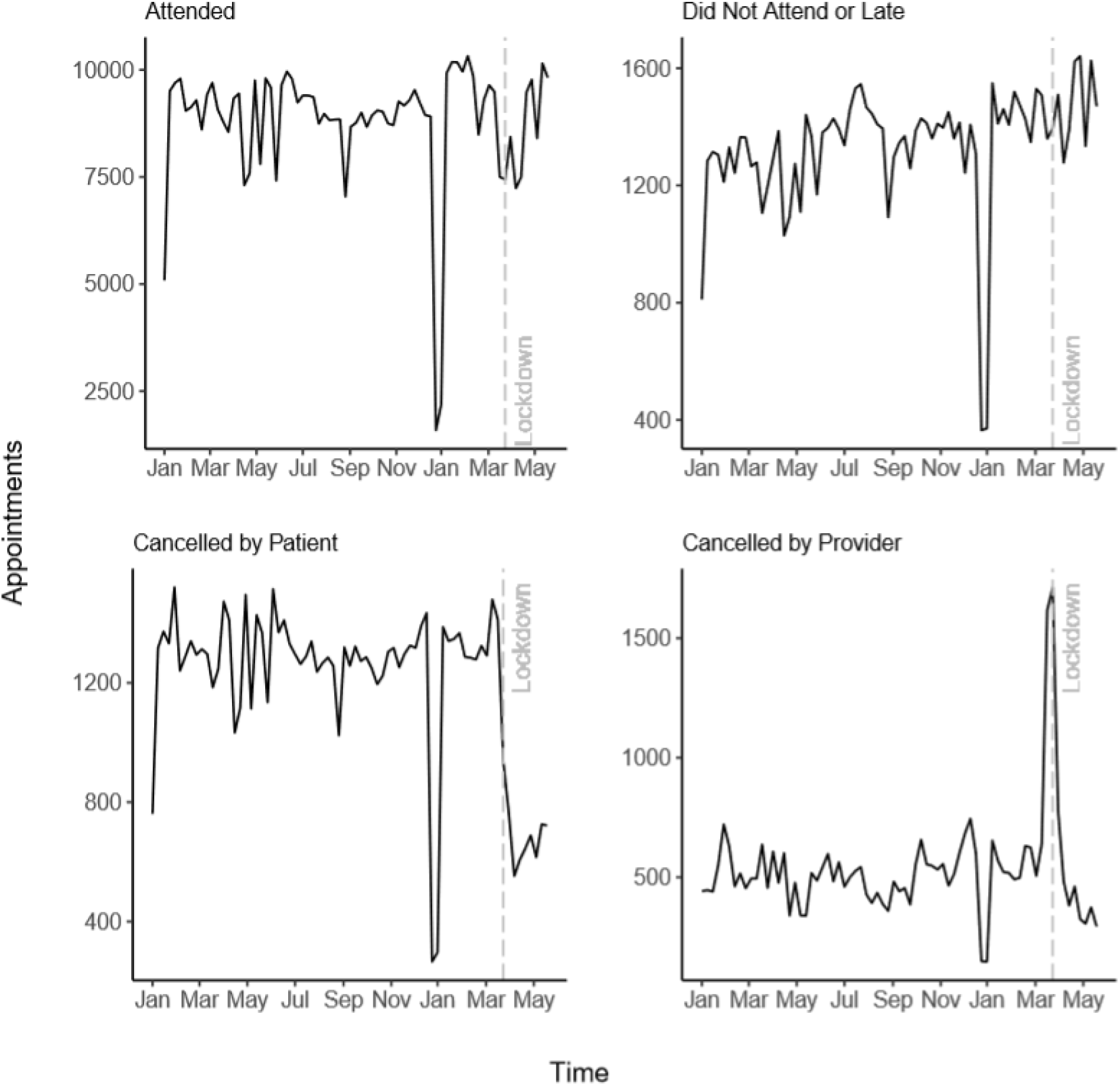
Weekly total appointments by attendance from 1st January 2019 to 24th May 2020

There was a brief, relatively small dip in attended appointments around lockdown; however, this did not appear to deviate strongly from attended appointments during the same time in 2019. There was a marginal increase in patients who Did Not Attend (DNA) appointments or attended too late to be seen after lockdown, which is greater than the DNA or late appointments at the same time in 2019. There was a large reduction in appointments cancelled by patients after lockdown, whereas there was a brief spike in appointments cancelled by providers around lockdown.

**Figure 7.**
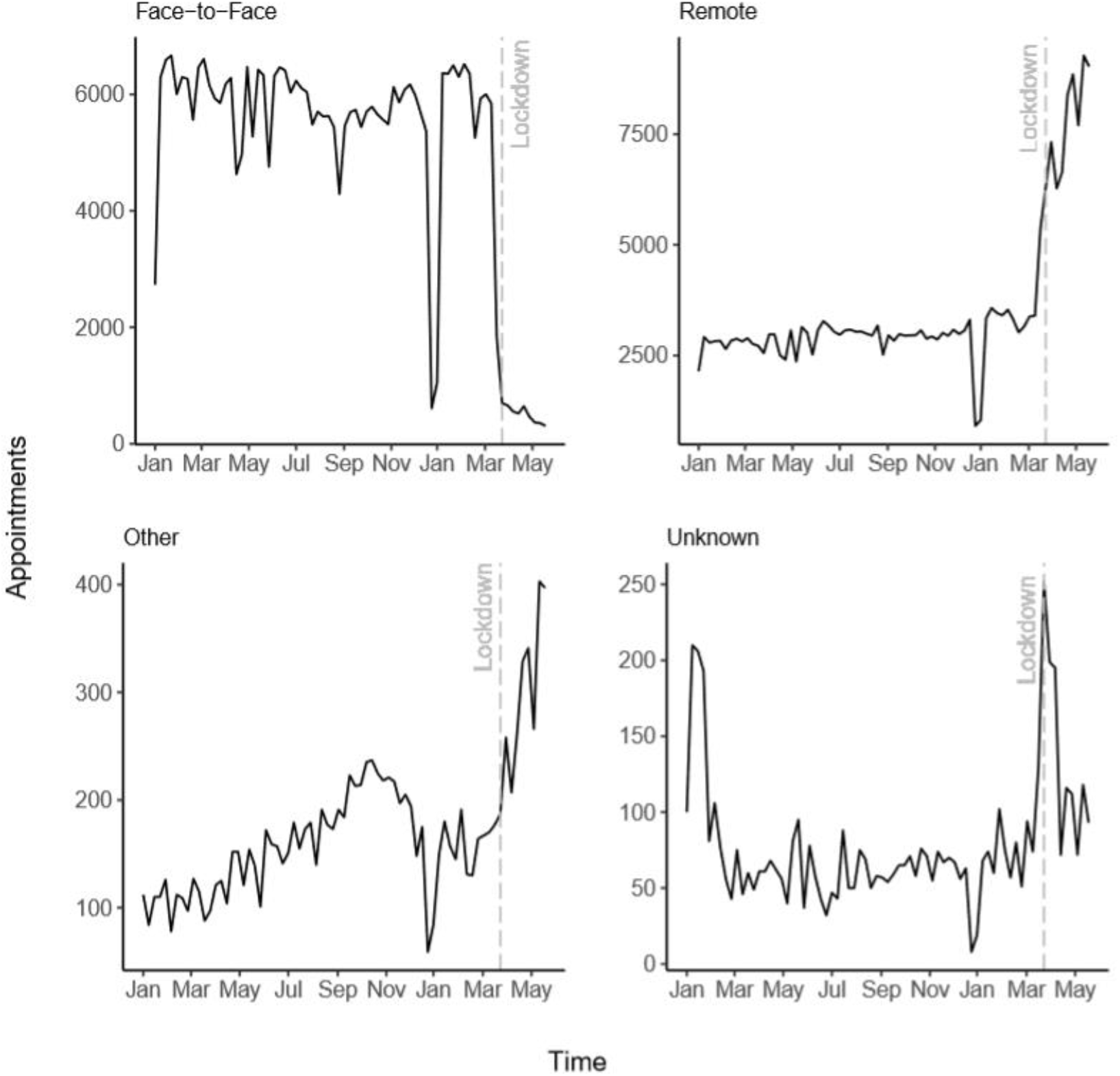
Weekly total attended appointments by consultation medium from 1st January 2019 to 24th May 2020

Out of all attended appointments, face-to-face consultations significantly reduced immediately after the lockdown was imposed, with remote appointments increasing. There was also an increase in appointments labelled *other* as well as a short spike in missing data for consultation medium. This may reflect a quick transition to different or novel consultation mediums that had not been previously used and thus not recordable in the patient recording system. An example of this might be the use of remote meeting software.

## DISCUSSION

Using electronic healthcare records, we examined the short-term impact of COVID-19 on primary care psychological therapy services in England, with regard to access and service provision. There was a clear drop in referrals to IAPT around the implementation of lockdown, resulting in approximately 55% fewer patients accessing services in the early weeks after the lockdown was imposed in the UK. There appeared to be a trend indicating faster increases in the number of referrals from BAME, especially Black, patients and patients living in urban areas once referrals started to increase again. Despite reductions in the number of people accessing services, it appears that the care of patients receiving treatment showed short-lived disruptions, with services quickly moving to provide remote consultations.

Overall, there was an average reduction of 55% in referrals in the 9 weeks after lockdown when compared to the same timeframe in 2019. This decline in referrals began approximately one week prior to lockdown and reached the maximum level three weeks after the lockdown was announced, with a 74% reduction of referrals in 2020 compared to the same time in 2019. This is consistent with the national trend observed through the monitoring of activity in the patient management software used by a majority of IAPT services, where referrals dropped by approximately 70% early after lockdown was announced (Eldridge, 2020). Patterns in the use of primary care psychological therapies are similar to those observed in other health services, such as a reduced number of patients accessing Accident and Emergency Departments and General Practice (Charlesworth, 2020; The Health Foundation, 2020). Although not returning to baseline, the number of referrals appeared to gradually increase again over time, with referrals being 24% lower at the end of May 2020 compared to 2019. Albeit slightly lower, a similar pattern is observed at a national level, where referrals were at 60% of the volumes observed prior to COVID-19 in July 2020 (Eldridge, 2020). Despite referral rates increasing again as the lockdown progressed, a large deficit in referred patients was observed. If the present research is extrapolated across England, with an assumed 1.69 million referrals per year, as observed in 2019–20, approximately 160,875 patients who may have normally been referred did not access mental health services in the 9 weeks after lockdown was imposed (NHS Digital, 2020). However, this deficit estimate is likely conservative – figures may be higher in the longer term as referrals had not returned to baseline towards the end of May and the proposed figure does not account for the annual increase in referrals (NHS Digital, 2020).

Self-referrals appeared to have recovered from the effects of lockdown most rapidly, which may have been facilitated through an increase in the availability and use of online referral systems (Eldridge, 2020). Online self-referrals to IAPT were 13% higher in July 2020 than pre-lockdown (Eldridge, 2020). We also observed differences in the referral rate by sociodemographic characteristics. Referrals from BAME groups appeared to be returning to baseline most rapidly after the initial observed decline. Referrals from patients with a Black background showed a particularly fast increase after the initial fall, reaching the highest number of weekly referrals observed in 2019/2020 towards the end of May 2020. It also appears that referrals after lockdown are primarily driven by people living in more densely populated areas. It is tentatively suggested that this could reflect a greater impact of COVID-19 on the mental health of BAME and urban populations, which would be consistent with emerging findings highlighting inequalities of COVID-19 (Elwell-Sutton, Deeney, & Stafford, 2020) and higher levels of depression and anxiety amongst BAME and urban populations throughout lockdown (Fancourt, Bu, Mak & Steptoe, 2020). However, population-based surveys have also identified higher sustained levels of depression and anxiety throughout lockdown amongst young adults and people from low-income households (Fancourt, Bu, Mak & Steptoe, 2020). In the present study, an increased demand for psychological therapy by these groups would be reflected by an increase in average IMD and a decrease in age at referral. However, we found no clear evidence that the average IMD or age of referrals to IAPT services changed as a result of the pandemic, suggesting there could potentially be a gap in access to mental health support amongst these populations.

There was an observed change in the clinical severity of referrals, with lower average depression and anxiety scores of incoming referrals prior to lockdown and an increase after the lockdown had been implemented in the UK. This increase remained somewhat stable for depression, however appeared to be returning to the annual average for anxiety towards mid-May 2020. Due to the observational nature of the data, it is difficult to discern whether increases in depression and anxiety resulted from a rise in symptoms amongst the general population or whether patients with more severe symptoms were accessing services to a greater degree after the lockdown was imposed. However, population-based surveys showed a decrease in both depression and anxiety from the start of lockdown until May (Fancourt, Bu, Mak & Steptoe, 2020). As such, the latter is suggested as more probable.

While there was an evident impact of COVID-19 lockdown on people accessing primary mental services, it appears that service delivery for patients already being treated pre-lockdown or starting treatment during lockdown saw limited disruption as services adapted quickly to new practices. This is consistent with national trends showing no dramatic fluctuations in clinician activity recorded by the IAPT patient management software, with the largest observed reduction in the number of appointments reaching approximately 24% (Eldridge, 2020). Services appeared to rapidly adapt, implementing infection control measures by switching to remote consultations almost exclusively. Mental health staff accounts mirror this, reporting rapid innovation with a particular emphasis on remote working (Johnson et al., 2020). Patterns in primary care appear similar to those observed in secondary care (Stewart, Martin & Broadbent, 2020).

### Strengths & Limitations

The present study is the first to examine a quantifiable impact of COVID-19 on primary care mental health services at scale, with data from a wide and diverse range of service providers across multiple geographic regions in England. Nonetheless, the dataset contains only a subset of service providers and may therefore not be nationally representative. There may be variation by service providers and regions that is not captured by the data used in the present study. However, the observed trends appear consistent with those detected by the monitoring of activity within IAPT service’s patient management software (Eldridge, 2020). Furthermore, it should be noted that the present research took a cross-sectional, observational approach using descriptive methods. As such, it is not possible to draw causal conclusions, and the capacity of estimating future impact is limited and remains speculative.

### Implications

A clear reduction of referrals took place during the early stages of COVID-19, producing approximately a 55% deficit in patients receiving mental healthcare. A concern may be that a backlog of patients has accumulated, which may cause future pressures on service providers to treat these patients in addition to an possible excess of patients who may seek mental health support as the long-term consequences of COVID-19 become more apparent. Given the faster increase in referrals after the initial drop from urban and BAME groups, compared to other groups, service providers catering to these populations may experience a particular surge in demand. This also serves as a reminder of the need for cultural competency in psychological therapy to meet the needs of all patients accessing services (Naz, Gregory, & Bahu, 2019). Periodical horizon scanning of the demography of patients accessing services may provide an avenue to assure that developments in cultural competency adequately reflect demographic changes.

While population-based surveys have observed that young adults and individuals from low-income households have had sustained higher levels of depression and anxiety throughout the pandemic, we found no strong evidence to suggest differences in age or deprivation of incoming referrals (Fancourt, Bu, Mak & Steptoe, 2020). This could begin to suggest a gap in access to mental health services for individuals who are particularly vulnerable to the mental health consequences of COVID-19 and warrants further investigation.

Despite access to mental health services being impacted by COVID-19, the data suggests that service providers rapidly adapted to the pandemic with the adoption of remote consultations. This shift likely provided essential continuity of care to patients in receipt of mental healthcare. Previous research suggests that remote Cognitive Behaviour Therapy is effective (Cuipers, Noma, Karyotaki, Cipriani & Furukawa, 2019: Linde et al., 2015) and may increase adherence (Mohr et al., 2012). However, approximately 40% of community and psychological therapy staff have reported difficulties with learning new technologies too quickly or without enough training and experiencing technical difficulties with remote consultation (Johnson et al., 2020). Furthermore, remote therapy may come at the cost of poorer maintenance after treatment (Mohr et al., 2012). As such, the effects of this rapid shift in working pattern on short- and long-term clinical outcomes remains to be determined, with a pressing need for future research.

### Conclusion

The present findings provide insight into the short-term impact of the COVID-19 pandemic on mental health services. Due to the observational nature of the data, results should be interpreted with caution; however, they have the potential to support the planning of clinical practice and public health policy as well as presenting avenues for future research. The long-term impact of COVID-19 on mental health services and mental health more generally remains to be determined as the delayed consequences, such as economic hardship, become more apparent.

## Data Availability

Data ownership lies with the NHS; the authors do not have permission to distribute data used in the present analysis. Data can be obtained by seeking relevant regulatory approvals.

## DECLARATIONS

### CRediT Author Statement

Clarissa Bauer-Staeb – Conceptualisation, Methodology, Writing – Original Draft, Formal Analysis, Visualisation.

Alice Davis – Conceptualisation, Methodology, Writing – Review & Editing.

Theresa Smith – Conceptualisation, Writing – Review & Editing.

Chris Eldridge – Resources, Writing – Review & Editing.

Wendy Wilsher – Data Curation.

David Betts – Resources, Writing – Review & Editing.

Emma Griffith – Writing – Review & Editing, Supervision.

Julian Faraway – Conceptualisation, Methodology, Writing – Review & Editing, Supervision.

Katherine Button – Conceptualisation, Methodology, Writing – Review & Editing, Supervision.

### Funding Statement

AD and TS were funded by Innovate UK (KTP #11105). The funders had no role in study design, data collection and analysis, decision to publish, or preparation of the manuscript.

### Conflicts of Interest

CBS, JF, KB, TS, AD, DB, WW, CE report no conflicts of interest. EG is a member of British Psychological Society, Division of Clinical Psychology, Digital Healthcare subcommittee and a coinvestigator on a grant from the British Psychological Society (“Developing Competencies for Digital Practice”).

### Ethical Approval

Due to the anonymous nature of the data, the present study was exempt from NHS Ethical Review. The project received ethical approval by the University of Bath (PREC: 19–015).

## Acknowledgments

We would like to thank all service providers and patients whose anonymous data was made available for the present research. We would further like to thank Mayur Bhatt (Avon and Wiltshire Mental Health Partnership NHS Trust) for helpful discussions regarding the implications of the present research.

## Notes

### Author Declarations

Due to the anonymous nature of the data, the present study was exempt from NHS Ethical Review. The project received ethical approval by the University of Bath (PREC: 19-015).

## REFERENCES

1 Mchugh, R. K., Whitton, S. W., Peckham, A. D., Welge, J. A., & Otto, M. W. (2013). Patient Preference for Psychological vs Pharmacologic Treatment of Psychiatric Disorders. The Journal of Clinical Psychiatry, 74(06), 595–602. doi:10.4088/jcp.12r07757

2 Clark, D. M., Canvin, L., Green, J., Layard, R., Pilling, S., & Janecka, M. (2018). Transparency about the outcomes of mental health services (IAPT approach): An analysis of public data. The Lancet, 391(10121), 679–686. doi:10.1016/s0140-6736(17)32133-5

3 NHS Digital. (2020, July 30). Psychological Therapies, Annual report on the use of IAPT services 2019–20. Retrieved August 5, 2020, from https://digital.nhs.uk/data-andinformation/publications/statistical/psychological-therapies-annual-reports-on-the-use-ofiapt-services/annual-report-2019-20

4 Holmes, E. A., O’connor, R. C., Perry, V. H., Tracey, I., Wessely, S., Arseneault, L., … Bullmore, E. (2020). Multidisciplinary research priorities for the COVID-19 pandemic: A call for action for mental health science. The Lancet Psychiatry, 7(6), 547–560. doi:10.1016/s2215-0366(20)30168-1

5 Fancourt, D., Steptoe, A., & Bu, F. (2020). Trajectories of depression and anxiety during enforced isolation due to COVID-19: longitudinal analyses of 59,318 adults in the UK with and without diagnosed mental illness. medRxiv. doi: https://doi.org/10.1101/2020.06.03.20120923.

6 Bu, F., Steptoe, A., & Fancourt, D. (2020). Loneliness during lockdown: trajectories and predictors during the COVID-19 pandemic in 35,712 adults in the UK. medRxiv. doi: https://doi.org/10.1101/2020.05.29.20116657

7 Frank, P., Iob, E., Steptoe, A., & Fancourt, D. (2020). Trajectories of depressive symptoms among vulnerable groups in the UK during the COVID-19 pandemic. medRxiv. doi: https://doi.org/10.1101/2020.06.09.20126300

8 Moorthy, A., & Sankar, T. K. (2020). Emerging public health challenge in UK: Perception and belief on increased COVID19 death among BAME healthcare workers. Journal of Public Health, Fdaa096. doi:10.1093/pubmed/fdaa096

9 Johnson, S., Dalton-Locke, C., San Juan, N. V., Foye, U., Oram, S., Papamichail, A., … & Rains, L. S. (2020). Impact on mental healthcare and on mental health service users of the COVID-19 pandemic: a mixed methods survey of UK mental healthcare staff. medRxiv. doi: https://doi.org/10.1101/2020.06.12.20129494

10 Stewart, R., Martin, E., & Broadbent, M. (2020). Mental health service activity during COVID-19 lockdown: South London and Maudsley data on working age community and home treatment team services and mortality from February to mid-May 2020. medRxiv. doi: https://doi.org/10.1101/2020.06.13.20130419

11 Kroenke, K., Spitzer, R. L., & Williams, J. B. (2001). The PHQ-9: Validity of a Brief Depression Severity Measure. Journal of General Internal Medicine, 16(9), 606–613. doi:10.1046/j.1525-1497.2001.016009606.x

12 Spitzer, R. L., Kroenke, K., Williams, J. B., & Löwe, B. (2006). A Brief Measure for Assessing Generalized Anxiety Disorder. Archives of Internal Medicine, 166(10), 1092. doi:10.1001/archinte.166.10.1092

13 Noble, S., McLennan, D., Noble, M., Plunkett, E., Gutacker, N., Silk, M., & Wright, G. (2019). The English indices of deprivation 2019. Hampshire: Office for National Statistics.

14 Park, N. (2020). Population estimates for the UK, England and Wales, Scotland and Northern Ireland, provisional: mid-2019. Hampshire: Office for National Statistics.

15 R Core Team (2013). R: A language and environment for statistical computing. R Foundation for Statistical Computing, Vienna, Austria. URL http://www.R-project.org/.

16 Eldridge, C. (2020, August 5–6). Covid-19: Trends in IAPT data [Conference presentation]. Digital Innovation in Mental Health, London, United Kingdom. Retrieved August 6, 2020, from https://www.youtube.com/watch?v=AtvO5ryicF8&feature=youtu.be

17 Charlesworth, A. (2020, May 28). Shock to the system: COVID-19’s long-term impact on the NHS. [Web blog post]. Retrieved July 23, 2020, from https://www.health.org.uk/news-and-comment/blogs/shock-to-the-system-covid-19slong-term-impact-on-the-nhs

18 The Health Foundation. Visits to A&E departments in England in April 2020 fell by 57% compared to last year. (2020, May 15). Retrieved July 14, 2020, from https://www.health.org.uk/news-and-comment/charts-and-infographics/visits-to-a-e-departments-in-england-in-april-2020-fell-by-57

19 Elwell-Sutton, T., Deeney, S., & Stafford, M. Emerging findings on the impact of COVID-19 on black and minority ethnic people. [Web blog post]. (2020, May 20). Retrieved July 14, 2020, from https://www.health.org.uk/news-and-comment/charts-and-infographics/emerging-findings-on-the-impact-of-covid-19-on-black-and-min

20 Fancourt, D., Bu, F., Mak, H., & Steptoe, A. (2020). Covid-19 Social Study Results Release 15. (Rep.). London: University College London. https://b6bdcb03-332c-4ff9-8b9d-28f9c957493a.filesusr.com/ugd/3d9db5_17cc74c304664db8ac9ea56e1dd301ae.pdf

21 Naz, S., Gregory, R., & Bahu, M. (2019). Addressing issues of race, ethnicity and culture in CBT to support therapists and service managers to deliver culturally competent therapy and reduce inequalities in mental health provision for BAME service users. The Cognitive Behaviour Therapist, 12. doi: https://doi.org/10.1017/S1754470X19000060

22 Cuijpers, P., Noma, H., Karyotaki, E., Cipriani, A., & Furukawa, T. A. (2019). Effectiveness and Acceptability of Cognitive Behavior Therapy Delivery Formats in Adults With Depression. JAMA Psychiatry, 76(7), 700. doi:10.1001/jamapsychiatry.2019.0268

23 Linde, K., Rücker, G., Sigterman, K., Jamil, S., Meissner, K., Schneider, A., & Kriston, L. (2015). Comparative effectiveness of psychological treatments for depressive disorders in primary care: Network meta-analysis. BMC Family Practice, 16(1). doi:10.1186/s12875-015-0314-x

24 Mohr, D. C., Ho, J., Duffecy, J., Reifler, D., Sokol, L., Burns, M. N., … Siddique, J. (2012). Effect of Telephone-Administered vs Face-to-face Cognitive Behavioral Therapy on Adherence to Therapy and Depression Outcomes Among Primary Care Patients. JAMA, 307(21). doi:10.1001/jama.2012.5588

